# Automated Contact Tracing: a game of big numbers in the time of COVID-19

**DOI:** 10.1101/2020.04.22.20071043

**Authors:** Hyunju Kim, Ayan Paul

## Abstract

One of the more widely advocated solutions for slowing down the spread of COVID-19 has been automated contact tracing. Since proximity data can be collected by personal mobile devices, the natural proposal has been to use this for automated contact tracing providing a major gain over a manual implementation. In this work, we study the characteristics of voluntary and automated contact tracing and its effectiveness for mapping the spread of a pandemic due to the spread of SARS-CoV-2. We highlight the infrastructure and social structures required for automated contact tracing to work. We display the vulnerabilities of the strategy to inadequate sampling of the population, which results in the inability to sufficiently determine significant contact with infected individuals. Of crucial importance will be the participation of a significant fraction of the population for which we derive a minimum threshold. We conclude that relying largely on automated contact tracing without population-wide participation to contain the spread of the SARS-CoV-2 pandemic can be counterproductive and allow the pandemic to spread unchecked. The simultaneous implementation of various mitigation methods along with automated contact tracing is necessary for reaching an optimal solution to contain the pandemic.

## Introduction

A relentless and damaging battle is being fought against the spread of COVID-19. While several countries have managed to significantly slow down its spread, severe measures have had to be taken to do so and at great cost to the economic and social well-being of the nations. It is still not certain when a significant control over the spread of SARS-CoV-2 can be attained. Recent projections propose surveillance for the next few years [1], with several measures that will need to be put in place to minimize the cost of the pandemic to humankind. Automated contact tracing is one of these measures.

Contact tracing has been observed to be effective in previous pandemics (or epidemics) like the Ebola virus outbreak in 2014-2015 [2]. This preemptive method allows for the containment of the pathogen by isolating potentially infected individuals that have been traced. Extensive studies of manual contact tracing were done during the previous outbreak of the Ebola virus [3-5], SARS-CoV and MERS-CoV [6]. More recently, mathematical models have been formulated to study contact tracing assuming the disease spread to be quantifiable by the SIR model [7]. However, the efficacy of automated contact tracing during the SARS-CoV-2 pandemic requires a more detailed examination given the distinct difference in the prevalence of this pandemic from the ones in the recent past and the different modes of transmission of the pathogen.

Manual contact tracing is not very effective against pathogens that spread like the influenza virus but is more effective for containing smallpox and SARS-CoV and partially effective in containing foot-and-mouth disease [8]. The viral shedding patterns of SARS-CoV and MERS-CoV are similar [9, 10] and show almost no pre-symptomatic transmission [11],^1^ while Ebola is known to be transmitted through the bodily fluids of infected individuals after the onset of symptoms [13]. On the other hand, influenza shows a significant rate of viral shedding in the pre-symptomatic stage [14]. The important transmission characteristics of SARS-CoV-2 that set it apart from other HCoV pathogens like SARS-CoV and MERS-CoV and from Ebola are:

- SARS-CoV-2 transmission is driven by pre-symptomatic spreading like the influenza virus [15-17].
- The pathogen can be transmitted through the air in high contamination regions and through contaminated dry surfaces for several days [15,18,19] leading to its high transmission rates. This brings about additional challenges when the disease cannot be contained within an isolated envelope of a healthcare system. While a similar spreading pattern is seen in SARS-CoV and MERS-CoV, this makes SARS-CoV-2 more easily transmittable than Ebola.
- The ACE2 binding of SARS-CoV-2 is estimated to be relatively stronger than SARS-CoV and might explain its observed spreading characteristics [20-22].

Similar to SARS-CoV the reproduction number, *R*_0_, for SARS-CoV-2 is estimated to be 2.2 - 2.7 [23-27].^2^ The dispersion parameter is estimated to also be similar to that of SARS-CoV (close to 0.1), which could be causing superspreading [26,29-31].

While, theoretically, automated contact tracing can be shown as an effective means of containing SARS-CoV-2 [30], factors such as long delays from symptom onset to isolation, fewer cases ascertained by contact tracing, and increasing pre-symptomatic transmission can significantly impact how effective contact tracing will be in practice. Defining significant contact as being within 2 meters and lasting for at least 15 minutes can result in the detection of more than 4 out of 5 secondary infections but at the cost of tracing 36 contacts per individual (95^th^ percentile: 0 - 182 contacts per individual) [32]. Changes to the definitions of the parameters can reduce the numbers traced. If the threshold of minimum contact time is increased to at least 4 hours of contact the spread of the pathogen cannot be controlled by automated contact tracing since many potentially infected contacts will escape detection. Detailed modeling of SARS-CoV-2 transmission shows that the pandemic can be sustained just by pre-symptomatic transmission and that automated contact tracing can be used to contain the spread of the pathogen if there are no significant delays to identifying and isolating infected individuals and their contacts [33].

Considering all the factors that make contact tracing a different game for SARS-CoV-2, in this paper we will examine in detail how much data and participation from the population will be needed to make automated contact tracing effective. This will give an estimate of the necessary scale of implementation of automated contact tracing and whether it will be feasible. The model that we build with factorized parameters will also allow for the estimation of the effects of various mitigation methods like the use of PPE in enhancing the efficacy of automated contact tracing which we discuss before the discussion section. In this work we address voluntary and automated contact tracing using proximity data alone excluding methods such as the use of CCTV, credit card information, logging of identities of individuals during vists to locations and travels, etc. that have been successfully used by many countries like Singapore [34], Taiwan [35], South Korea [36] and China [33] for contact tracing.

## Contact tracing for COVID-19

To begin with, we consider a disease that spreads only in the symptomatic stage. The infected individuals can spread the disease to their contacts before they are isolated and to medical workers after they are isolated with varying probabilities. Of significance here is that after the initial period of ignorance of the population about a rising pandemic, infected individuals will be isolated with higher efficiency (even with manual contact tracing) resulting in the curtailment of the spread of the pathogen. How is contact tracing more effective in such diseases? Since the mobility of the infected individual usually sees a decline after the onset of symptoms, the number of contacts at risk become limited to nearest neighbors and possibly next-to-nearest neighbors in the contact space. This allows the implementation of a manual contact tracing algorithm that identifies these neighbors and isolates or tests them as suggested in reference [8]. This was seen to be effective during the Ebola, MERS-CoV and SARS-CoV outbreaks.

However, the spreading of SARS-CoV-2 follows a very different pattern. With the prevalence of spreading of infection through pre-symptomatic and subclinical hosts, the number of individuals that might need to be traced can be very large. This has led to the belief that automated contact tracing in a wider gamut should be implemented. Most of the proposed solutions [30,32,33] require the use of historical proximity data to trace contacts. In the context of COVID-19, there are some obvious pitfalls in the algorithm:

- It is estimated that about 86% (95% CI: [82% - 90%]) of the infected cases in China were undocumented prior to the travel ban on the 23^rd^ of January 2020 generating 79% of the documented infections [23]. A large number of these undocumented cases experienced mild, limited or no symptoms and can hence go unrecognized. Similar results were reported by other studies [37,38]. It is not possible to trace all the contacts of these individuals since they will be partially reported leading to incomplete coverage of contact tracing.
- While it is assumed that the SARS-CoV-2 spreads within a proximity radius of *r*_0_ (assumed to be 2 meters), not much is known about the probability of transmission, *p_t_* when two individuals come within this domain of contact for a minimum contact time *t*_0_. Assuming *p_t_* to be large will lead to an unreasonably large estimate of the number of potential contacts in a crowded region like supermarkets, which remain open even during the period of social distancing. On the other hand, assuming pt to be small will underestimate the number of infected contacts, especially because there might be other modes of transmission of SARS-CoV-2 that are not being considered. By definition *p_t_* depends on the dynamics of disease transmission when a healthy individual comes in significant contact with a sick individual. Moreover, pt is not constant over *r*_0_ and also varies with the stage of infection the infected individual is at [17]. Several other factors contribute to the value of pt in addition to the contagiousness of the disease including, but not limited to, the use of PPE, public awareness of the disease, etc.

The first pitfall can be alleviated by increasing the testing rate of individuals for viral RNA in the hope that a larger fraction of the asymptomatic or mildly symptomatic carriers can be traced. Increasing awareness can also help. The second pitfall can be alleviated when more detailed knowledge of the spread of SARS-CoV-2 is available and with the help of simulation of the spread of the disease in a population. For the rest of the work, we will assume pt to be a variable and *r*_0_ to be fixed to 2 meters [32].

The real-world applicability of automated contact tracing requires the examination of the effects of finite sampling of the population. The assumption that we are working with is that enrollment in automated contact tracing will be voluntary and individuals remain free to do one of the following:

- Choose not to enroll in the program by either not using the application or the devices needed for tracing, including discontinuity in participation.
- Choose not to report on their health condition which is assumed to be voluntary.

Both types of occurrences have an effect of reducing the efficacy of automated contact tracing but in slightly different manners. In the first case, not subscribing to the service would not only remove an individual from the pool that is being notified, but it also removes them from the pool of individuals that are reporting. In the second case only the latter happens.

## Modeling automated contact tracing

Since, in automated contact tracing, a significant contact has to be less than r0 distance away for time *t*_0_, we describe every individual by a circle with a radius of *r*_0_=2 which we shall call the cross-section of the individual. The cross-section is chosen such that any overlap between two cross-sections can be taken as a significant contact between the two respective individuals. Temporally, the cross-sections have to overlap for a time t0 which is the threshold interaction time that is assumed critical for an individual to infect another by proximity. For the sake of simplicity and without any loss of generality of our argument we can assume that the probability of getting infected, *p_t_*, is independent of the degree of overlap of the cross-sections and for any time *t* > *t*_0_ as is done normally in automated contact tracing.

Figure 1 gives a depiction of what automated contact tracing would be for a group of individuals. In the left-most panel, *B* and *C* are in contact with *A* at *t* = 0 but not with each other. *D* is isolated from all of them. After a period of time *t* < *t*_0_, *B* is isolated but *C* stays in contact with *A*. Then at time *t* = *t*_0_+*ε*, where *ε* ≪ *t*_0_, we see that *C* is still in contact with *A*, *B* remains isolated and *E* has come in contact with both *A* and *C*. Using the methods of automated contact tracing, if *A* reports sick at a future time, *C* will be deemed as having had significant contact with *A*. *E* might also be deemed as such depending on how long he maintains proximity with *A*, but the proximity of *E* with C need not be counted even if E spends *t* > *t*_0_ in contact with *C* (if only primary contacts are traced) unless *C* reports sick too.

**Figure 1.**
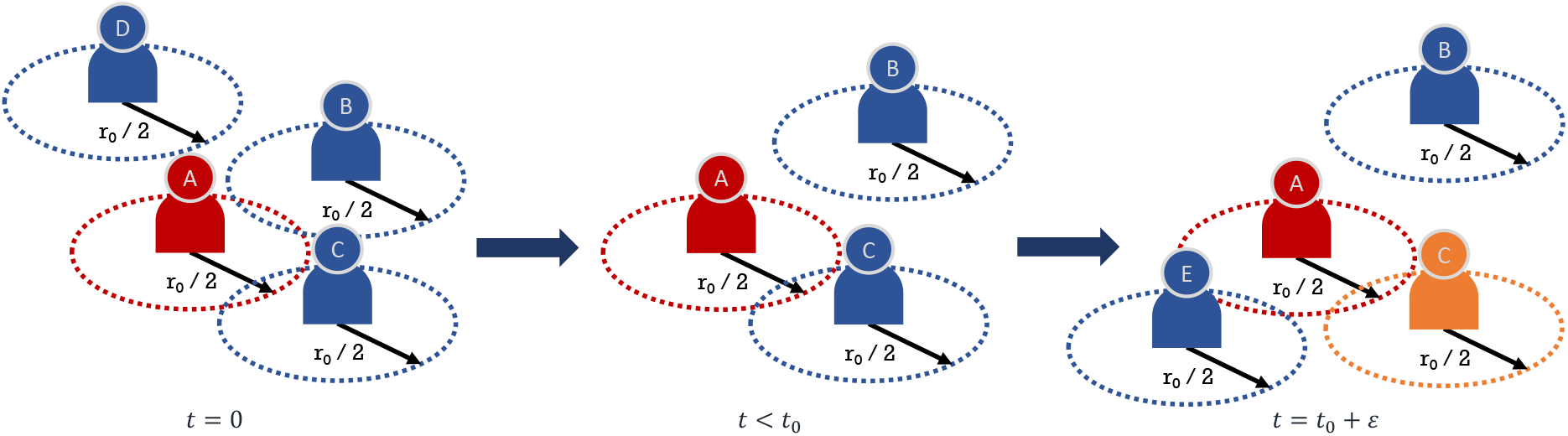
A depiction of automated contact tracing. The cross-section is denoted by the dashed circle and is of radius *r*_0_=2. Interactions occur from *t* = 0 to *t* = *t*_0_ + *ε* where *ε* ≪ *t*_0_. A will be confirmed as COVID-19 positive in the future and *C* will be notified having come in contact with *A*. *E* might be notified if *E* stays in contact with *A* for a time period greater than *t*_0_.

This method of automated contact tracing will work as long as *A* and *C* (and possibly *E*) are enrolled in the service even if *B* and *D* are not. However, *D* is completely isolated and by remaining so for a long time is observing social distancing from any other individual. B is representative of an individual who observes partial social distancing. Hence, for *D* this service is not necessary and for *B* it is of limited value. If *C* is not enrolled in the service *C* will never get notified if *A* gets sick. *C* might fall sick or become an asymptomatic carrier and continue contaminating others. If *A* does not enroll in this service then *C* never gets notified leading to the same conclusions but *E* might get notified if *C* declares sick and *E* is enrolled in the service.

An estimated 45% of virus transmission occurs in the pre-symptomatic phase of an infected individual [33]. From both Hellewell et. al. [30] and Ferretti et. al. [33] it is seen that 60% - 80% of the contacts need to be successfully traced and quarantined instantly to contain the outbreak over a period of time.Prevalence of subclinical infections of SARS-CoV-2 further reduces the effectiveness of contact tracing. With automated contact tracing using a definition of *r*_0_ = 2 meters and *t*_0_ *=* 15 minutes more than 80% of the cases can be traced [32] if every infected case is reported. In what follows we create a simplified model of automated contact tracing to deduce the minimum fraction of the population that needs to enroll in the program for it to be effective.

- Let *N* be the number of individuals in a population and *f_i_* the fraction of the population that is infected, regardless of whether they know it or not. Therefore, the true number of infected individuals is *f_i_N*.
- If testing is conducted only when mild or severe symptoms are seen (i.e. excluding testing of asymptomatic cases), the number of confirmed cases is *r_c_f_i_N* with *r_c_* being the fraction of the infected that will be confirmed as infected by testing.
- We define *f_e_* as the fraction of the population that is enrolled for automated contact tracing and *f_c_* as the fraction of the users that will confirm that they have been diagnosed positive. Hence, the number of individuals that have tested positive, are using automated contact tracing and will confirm that they are sick is *f_c_f_e_r_c_f_i_N*.
- We define *a_c_* as the average number of contacts per person in the period of time t_0_ who are at risk of being infected due to proximity with a sick individual and is assumed to be greater than 0.^3^

Since only *f_e_* fraction of contacts are using the service, we can estimate the number of individuals that can be traced as *f_c_f_e_r_c_f_i_N* × *a_c_* × *f_e_*.

To compute the number of individuals that need to be quarantined or isolated since they are now at risk of being infected from coming in contact with a sick person, we define the following.

- Since *p_t_* is defined as the probability of transmission of infection within the proximity radius *r*_0_ being exposed for a time greater than *t*_0_, the number of individuals at risk is, at most *p_t_f_i_Na_c_*, i.e., *p_t_* × number of contacts of the group of infected individuals.
- Finally, we define *f_T_* as the fraction of the individuals at risk of being infected that needs to be successfully quarantined to quell the spread of the pathogen.

Therefore, the number of individuals that should be quarantined is *f_T_p_t_f_i_Na_c_*. For automated contact tracing to work effectively, we have,

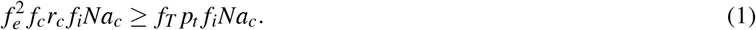

### The game of big numbers

Eq. (1) simply states that the number of individuals that can be notified by automated contact tracing (on the left-hand side) has to be greater than or equal to the number of individuals who need to be notified (on the right-hand side). Note that *a_c_*, the average number of contacts, drops out of the inequality and hence, the inequality is independent of the population density of the region since eq. (1) is in terms of fraction of the population and not the absolute number of individuals. This simply implies that in a region of denser population a larger number of people need to be contacted and quarantined but leaves *f_e_* independent of the population density. Since the right-hand side is the minimum fraction of the population that needs to be traced we arrive at:

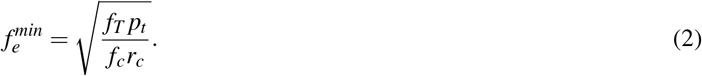

The fraction 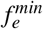 is the minimum fraction of the population that needs to be enrolled in automated contact tracing for it to be effective as a means of slowing down the spread of the pandemic. In eq. (2), *p_t_* depends on the spreading dynamics of the pathogen determined by individual-to-individual interactions and, therefore, also depends on the mitigating measures taken at both the population level and the individual level. Naively, in automated contact tracing, *p_t_* is taken as one if the contact has lasted for over time *t*_0_ with the subjects being less than *r*_0_ apart. This can be reduced by use of PPE or other mitigation methods as we discuss later. The parameter *f_T_* depends on the disease spreading dynamics and can be estimated from modeling the disease spreading amongst a population [33]. According to this reference, the slower the response to the identification of contact at risk higher is *f_T_* for the same reduction rate of the reproduction number. We assume that identification of contact at risk takes less than a day in automated contact tracing. The parameter *r_c_* is governed by the ability to identify infected individuals through testing and depends on the protocols of the testing program and its coverage. On the other hand, *f_c_* is determined solely by the degree to which individuals are willing and able to confirm that they have been tested positive.

Let us examine the limit *p_t_* = *f_c_* = *r_c_* = 1. This is the limit where every significant contact is assumed to be at risk, everyone who is enrolled in the automated contact tracing program reports sick when tested positive and every sick individual can be successfully identified by testing. Then we arrive at the relation 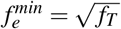 (blue dotted line in the third from left panel of figure 2). Since *f_T_* is the fraction of contacts that need to be successfully isolated, it can be extracted from the abscissa of fig. 3 of ref. [33]. For example, if 100% of the infected cases can be isolated, then for a change in the epidemic growth rate by -0.1, one needs *f_T_* ~ 60%. Hence 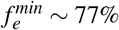. It is intuitive that 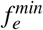 scales as the squareroot of *f_T_* since both the infected and the contact at risk need to be enrolled and the probability that each are enrolled is *f_e_* leading to 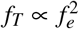. It gives the threshold which 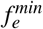 cannot exceed for any given *f_T_*.

Lastly, we define the effectiveness of the automated contact tracing, *η*, as the ratio of the actual number of individuals that will be notified (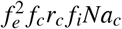) to the minimum number of individuals that should be notified to quell the spread of the disease (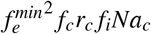) and get:

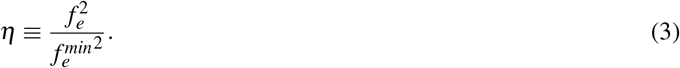

**Figure 2.**
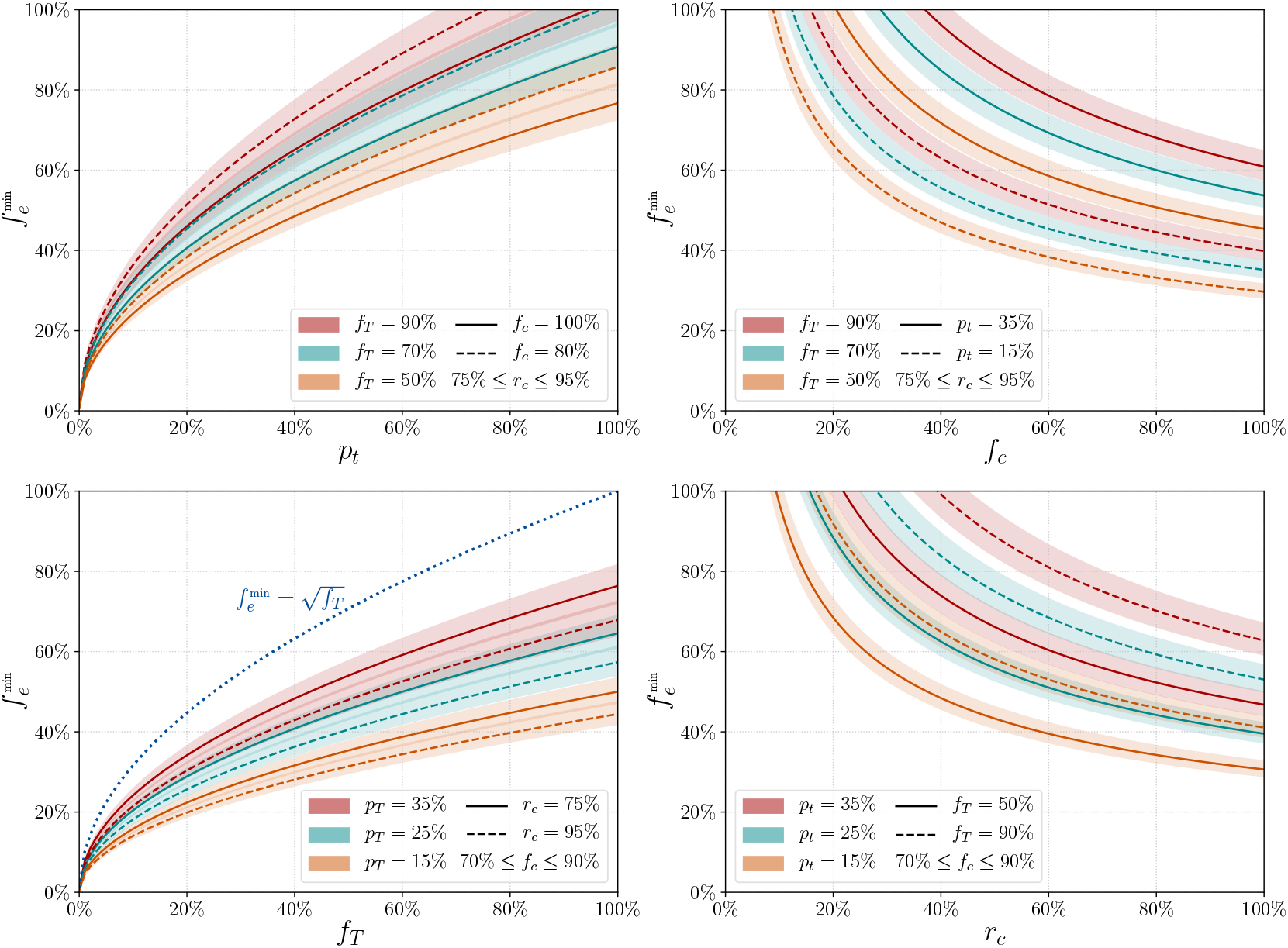
Percentage of the population that needs to be enrolled (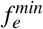) for automated contact tracing to be successful. Starting from the left, the solid and dashed lines represent *f_c_* = 100%, 80% respectively for the first panel, *p_t_* = 35%;15% for the second panel, *r_c_* = 75%;95% for the third panel and *f_T_* = 50%, 90% for the fourth panel. For the left two panels the fraction of truly infected individuals that will be confirmed as sick, *r_c_* is varied between 75%, 95%. For the right two panels the fraction of people who will confirm they are sick if they are enrolled, *f_c_* is varied between 70%, 90%. Three cases for the minimum fraction of the individuals at risk that need to be traced are considered with *f_T_* = 50%, 70%, 90% in orange, green and red respectively in the left two panels and similarly, three cases are considered for *p_t_* = 15%, 25%, 35% in the right two panels. The blue dotted line in the third panel from the left gives the threshold variation of 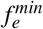 with *f_T_* when all other parameters are set to 1. The y axes are identical for all panels. See text for more details.

Figure 2 depicts how the fraction of the population that needs to be enrolled for the automated contact tracing program to be successful (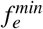) varies with the four factorized parameters. In the left-most panel of figure 2, we show the minimum percentage of the population that needs to be enrolled in automated contact tracing 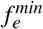 (in %) versus the transmission probability *p_t_*. We consider two values for *f_c_* = 80%, 100%, the fraction of individuals who test positive and will confirm their symptoms to trigger automated contact tracing, by the solid and dashed lines respectively. The solid and dashed lines represent *p_t_* = 15%, 25% respectively. The bands are generated by varying the fraction of sick individuals that can be confirmed as sick by testing, *r_c_*, between 75% and 90%. The other panels show the variation of 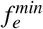 with *f_c_*,*f_T_* and *r_c_*.

If we take a closer look at eq. (1) and the left-most panel of figure 2 we see that even with a modest probability of transmission *p_t_* (e.g. about 30%) quite a large fraction of the population (about 40% - 60%) needs to be enrolled in automated contact tracing even when we assume almost all of them will be actively participating in confirming when they get infected. Assuming all the traced contacts within radius r0 lasting for more than t0 period of time are going to be infected is equivalent to stating *p_t_* = 100%. From the panel on the right, we can see how a fall in the fraction of individuals that confirm that they are sick, *f_c_*, can increase 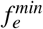. Even with quite low values of *p_t_* nearly half the population needs to be enrolled in automated contact tracing.

Let us try to understand why the effectiveness of automated contact tracing seems to drop so drastically with the enrollment fraction *f_e_*. From the left-hand side of eq. (1) we see that the effectiveness of automated contact tracing drops as 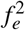. We see that *η* drops to 64% when 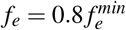 and 25% when 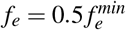. This non-linearity exists because *f_e_* not only reduces the number of sick individuals who can report their status but also the number of individuals who can receive a notification that they have come in contact with a sick person. The primary reason behind this is the fact that the automated contact tracing depends on voluntary participation whereas manual contact tracing or the use of CCTV, credit card information or identity logging at visited location to trace contact are not voluntary in their current form of implementation^4^.

Furthermore, as seen in figure 2, when the percentage of sick individuals who report that they are sick, *f_c_*, is lower than 100%, automated contact tracing becomes even less effective. In addition, the percentage of cases that can actually be detected, *r_c_*, will realistically be less than 100% for SARS-CoV-2 because of the prevalence of subclinical cases that will escape detection and other clinical factors.

In our analysis, we have inclined towards an optimistic picture of the spread of SAR-CoV-2. We have considered only spreading due to proximity and not considered other means of spreading like contaminated surfaces and aerosol that are common for SARS-CoV-2 [15, 18, 19] and can increase *p_t_*. In figure 2 we have taken a minimum *r_c_* of 75% when this can be even lower if widespread testing is not conducted to identify subclinical cases that can go undetected. We have also neglected the requirement for tracing secondary or tertiary contacts. In addition, we have also ignored events where a large number of individuals are infected in very a crowded location like public events for which thresholds like *r*_0_ and *t*_0_ need to be modified. Despite this optimistic picture, our analysis shows that a majority of the population has to enroll and actively participate in automated contact tracing for the measure to work in the absence of active social distancing measures.

We have not addressed the sociological aspect of selection bias in the enrollment process. Diversity in socio-economic conditions, awareness of technology and willingness to participate in a community effort will create variation in representations amongst the population. This can lead to the most vulnerable in society getting the least benefit from the implementation of automated contact tracing. Addressing the challenges of implementing automated contact tracing in developing nations where the necessary technologies might not be accessible to a large fraction of the population lies beyond the scope of this work.

### Assisted contact tracing

The necessary scale of implementation of automated contact tracing appears to be too large for it to be considered an effective measure to slow down the ongoing pandemic. For automated contact tracing to be a viable option, 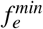 has to be as low as possible. To achieve this either the product *f_T_p_t_* needs to be decreased or the product *f_c_r_c_* needs to be increased as seen from eq. (2).

- Both *f_T_* and *p_t_* depend on the dynamics of the disease spreading amongst humans. The fraction of traced cases that need to be quarantined to stop the spread of the disease *f_T_* can be reduced by extensive monitoring of the disease to make sure sick cases are isolated as soon as possible and their contacts are traced. Even a day or two of delays can increase *f_T_* making automated contact tracing ineffective [33].
- Variations in *p_t_* can be caused by several factors some of which are controllable. Since *p_t_* depends on the contagiousness of the disease and any protective measures taken against the spread of the infection, *p_t_* can be reduced by measures of limited social distancing, the use of PPE and raising public awareness about the contagiousness of COVID-19. This can pose a significant challenge in densely populated regions and regions with poor living conditions and might lead to the breakdown of the applicability of automated contact tracing.
- *f_c_* is somewhat more difficult to control assuming the reporting of those who are confirmed sick is voluntary. This can only be increased by increasing the population’s willingness to contribute to automated contact tracing.
- *r_c_* is the parameter that is least under control since without very large-scale testing, asymptomatic and mildly symptomatic cases will be difficult to find. This is especially true if the infection can spread by means other than proximity alone as might be the case for SARS-CoV-2 [15,18,19].

Thus we see that a combination of several measures along with a large participation of the population in contact tracing would be the optimal solution for avoiding extensive population-wide social distancing measures and reducing the cost to the economy and well-being of a nation and also allow for greater freedom of movement during a pandemic.

## Discussion

We have shown that in real-world scenarios, automated contact tracing alone cannot contain a pandemic driven by a pathogen like the SARS-CoV-2. Advocating it as such can lead to an exasperating the spread of the pathogen. The primary reasons why such a strategy will not work as effectively as projected for SARS-CoV-2 is because of a large degree of spreading from pre-symptomatic and subclinical hosts, and the rapidity with which the virus spreads through proximity alone if no additional measures are taken to mitigate the spread. All of these conjugated with the vulnerability of automated contact tracing to insufficient sampling due to limited participation amongst the population and possibly incomplete reporting of sick cases will lead to reduction in the efficacy of automated contact tracing. A small fraction of the population being infected with SARS-CoV-2 can quickly lead to a majority of the population being needed to participate in the program.

We put together all the factors of concern and show that they follow a simple relationship. We further discussed how factors like the transmission probability *p_t_* should be reduced and the fraction of infected individuals that test positive, *r_c_*, should be increased to assist in reducing the burden on automated contact tracing while keeping the entire process voluntary. The strength of our model lies in the fat that we factorize the various parameters that separately contribute to the efficacy of automated contact tracing. This allows for individually addressing each parameter through improved clinical intervention, logistics, mitigation strategies and public awareness of automated contact tracing to increase adoption of the method. While our focus in this paper was to address the feasibility of automated contact tracing for containing the spread of SARS-CoV-2, eq. (2) can be applied for using automated contact tracing to contain other pathogens too. Our analysis is also independent of the methods of implementation of automated contact tracing and the definitions of *r*_0_ and *t*_0_. Therefore, our approach is quite general.

During the final stages of this work, a similar result was reached by the authors of [39] using a branching process model and arguments from statistical mechanics. They reached a similar conclusion as we do in our paper showing that nearly 75% to 95% of the population need to participate in automated contact tracing for it to be effective. The results in their work corresponds to ours when *p_t_ = f_c_ = r_c_ =* 1 or 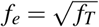.

The trust in contact tracing stems from the effectiveness with which it was used to contain pathogens like Ebola, SARS-CoV and MERS-CoV. However, the dynamics of the spread of SARS-CoV-2 is very different from these pathogens. Hence, the effectiveness of contact tracing in stopping the spread of these pathogens should not be seen as a validation of the effectiveness of automated contact tracing for SARS-CoV-2. To make automated contact tracing work, a majority of the population has to enroll for this service and actively participate in it. If this cannot be established then other measures of mitigating the spread of SARS-CoV-2 should be implemented in addition. As can be seen by the success of several nations in containing the spread of COVID-19, only a judicious combination of contact tracing with measures such as partial social distancing, wide use of PPE and dissemination of information about the disease can prove to be effective in slowing down the spread of the ongoing pandemic.

## Data Availability

All data necessary to reproduce our results are in the paper or the references therein.

## Acknowledgements

This work was partially supported by a grant from John Templeton Foundation. The opinions/conclusions presented in this publication are those of the author(s) and do not necessarily reflect the views of John Templeton Foundation. We would like to thank Paul Davies, Christophe Grojean, Luca Silvestrini and Melinda Varga for comments and suggestion.

## Footnotes

1 One study suggested that MERS-CoV can be transmitted before the onset of symptoms [12].

2 Much higher reproductive rates have also been estimated with data from Wuhan, China [28]

3 Here we make a simplifying assumption that the disease has spread to only a small fraction of the population and the probability of a single healthy person to meet two sick individuals within the proximity radius in a period of 14 days and be in contact with them for *t* ≥ *t*_0_ is negligibly small in general. There will be outliers depending on the habits of individuals but we can neglect them for this analysis.

4 These effectively makes f close to 100% for both those who have been diagnosed as infected and their contacts.

